# Is influenza B/Yamagata extinct and what public health implications could this have? An updated literature review and comprehensive assessment of global surveillance databases

**DOI:** 10.1101/2023.09.25.23296068

**Authors:** Saverio Caini, Adam Meijer, Marta C. Nunes, Laetitia Henaff, Malaika Zounon, Bronke Boudewijns, Marco Del Riccio, John Paget

## Abstract

**Introduction:** Early after the start of the COVID-19 pandemic, a major drop in the number of influenza B/Yamagata detections was observed globally. Given the potential public health implications, particularly with regards to influenza vaccination, we conducted a systematic review of influenza B/Yamagata virus circulation data from multiple complementary sources of information.

**Methods:** We searched articles published until 20^th^ March 2023 in PubMed and EMBASE; examined epidemiological and virological influenza data for 2020-2023 contained in the publicly available WHO-FluNet and GISAID (Global Initiative on Sharing All Influenza Data) global databases, or collected by the multi-national Global Influenza Hospital Surveillance Network (GIHSN) study; and looked for influenza data in the webpages of respiratory viruses surveillance systems from countries worldwide.

**Results:** Highly consistent findings were found across all sources of information, with a progressive decline of influenza B/Yamagata detections from 2020 onwards across all world regions, both in absolute terms (total number of cases), the positivity rate, and as a fraction of influenza B detections. Isolated influenza B/Yamagata cases continue to be sporadically reported, and these are typically vaccine-derived, mistaken data entries or under investigation.

**Discussion:** While it is still too early to conclude that B/Yamagata is (or will soon become) extinct, the current epidemiological and virological data call for a rapid response in terms of influenza prevention practices, particularly regarding the formulation of influenza vaccines. The current epidemiological situation is unprecedented in recent decades, underlying the importance of continuously and carefully monitoring the circulation of influenza viruses (as well as SARS-CoV-2 and the other respiratory viruses) in the coming years.

## Introduction

The emergence of the severe acute respiratory syndrome coronavirus 2 (SARS-CoV-2) and the resulting coronavirus disease (COVID-19) pandemic, including the public health measures that were put in place to contain the spread of the virus, caused major disruptions to the circulation of influenza [**Dhanasekaran 2022**] and most other respiratory viruses. Early in the course of the COVID-19 pandemic, the circulation of seasonal influenza viruses was drastically reduced, with most countries around the world reporting significant declines in influenza positivity rates (and, in some cases, only sporadic detections or even none at all) over the following usual epidemic periods [**Olsen 2020**]. During 2022, influenza viruses gradually resumed circulation [**Merced-Morales 2022**], although it is still to be determined whether the pre-2020 pattern of circulation (e.g. typical timing of start, peak, and end of seasonal epidemics) and burden of disease will be restored or instead changes will persist due to the co-circulation of influenza and SARS-CoV-2.

While much uncertainty remains and predictions are difficult to make, one aspect that quickly came to the attention of the scientific community was the strong decrease in the number of detections of influenza B/Yamagata cases. In the years prior to the emergence of SARS-CoV-2, the B/Victoria and B/Yamagata virus lineages tended to circulate simultaneously globally, with no clear or consistent pattern in the way the two lineages alternated from one season to another (including mixed seasons) [**Puzelli 2019**]. Interestingly, B/Yamagata viruses were responsible for a larger proportion of influenza B infections than B/Victoria globally during 2012-2017, but in the last two years prior to the COVID-19 pandemic (i.e. after the major outbreak of influenza B/Yamagata in 2017-2018 in large parts of the world), the B/Victoria lineage had become largely prevalent, and the B/Yamagata-to-B/Victoria ratio (calculated out of all influenza B cases that were characterized) dropped to 1/4.5 in 2018 and 1/19.3 in 2019 [**Zanobini 2022**]. Early in the course of the COVID-19 pandemic, and even after influenza viruses started circulating again in 2021, B/Yamagata lineage viruses were detected in only a very few countries globally according to previous reports [**Koutsakos 2021, Dhanasekaran 2022, Paget 2022**], and the question arose as to whether B/Yamagata lineage viruses had gone extinct (or pushed close to extinction) by the pandemic.

It is premature to definitely declare the extinction of the B/Yamagata lineage; as viral circulation might be at low levels, sub-threshold compared to the capabilities of existing surveillance systems or circulating in regions of the world currently not well covered by surveillance systems (e.g. West Africa), and there remains the possibility of a resurgence of the B/Yamagata lineage in the future. Indeed, it is worth mentioning that during much of the 1990s the B/Victoria lineage did not circulate in most regions of the world and was primarily confined to East Asia, before spreading globally again in the early 2000s [**Xu 2015**]. The current epidemiological situation holds significant implications for public health, particularly regarding influenza vaccination strategies [**Paget 2022, Dhanasekaran 2022**]. Since quadrivalent influenza vaccines (QIVs) contain virus strains from both B lineages, the question arises as to whether it is appropriate to recommend a vaccine targeting a virus that is currently not circulating (regardless of cost-effectiveness considerations and potential hesitancy among the population). Moreover, there is a concern that the mechanistic attenuated replication of the B/Yamagata strains contained in the live attenuated influenza vaccine (LAIV), primarily administered to children (e.g. UK, US and EU regions), could theoretically lead to reintroduction of the haemagglutinin (HA) (and neuraminidase (NA)) genes of a potentially extinct virus through reassortment with a coinfecting wildtype B/Victoria lineage virus [**Dudas 2015**]. Importantly, LAIV, when shedding, retains its cold-adapted nature and secondary infections after vaccine-derived influenza B/Yamagata infections are unlikely and indeed have not been shown to date [**Vesikari 2006**]. Continuous monitoring of viral circulation is therefore extraordinarily important so that influenza prevention policies, especially those related to vaccine formulation and composition, can be adapted and tailored to the current epidemiological landscape.

Here we aimed to study the circulation of B/Yamagata influenza viruses from 2020 onwards, in order to understand whether these viruses are extinct (or could become extinct soon) and discuss what public health implications this could have. For this purpose, we conducted a comprehensive and systematic review of multiple data sources pertaining to the circulation of B/Yamagata influenza viruses. This updated review builds upon prior assessments while aiming to have a higher degree of comprehensiveness than any previously published. To achieve this, our search has been extended from global databases for influenza virological surveillance (e.g. WHO-FluNet and GISAID) [**Paget 2022**], to include another surveillance database (GISHN), biomedical literature databases (PubMed and EMBASE), as well as the webpages of national surveillance systems around the world.

## Methods

### Systematic literature review

On March 20^th^, 2023, we searched PubMed and EMBASE using the following search string: ((influenza OR flu) AND (Yamagata OR Victoria OR lineage OR Vic OR Yam)) OR (“influenza b”). All articles published from January 1^st^, 2020 onwards were subjected to a multi-step selection process to evaluate their eligibility for inclusion in the literature review. The selection procedure consisted of an initial screening-out step in which two investigators (SC and BB) independently discarded irrelevant articles based on their title and abstract, and a second step in which all the articles retained after the first step by at least one of the two investigators were read in full text in order to evaluate their eligibility. We did not place any language restriction as long as an English title and abstract were available for the first step of the selection process. Finally, we included articles focusing on influenza in which the collection of respiratory specimens for influenza testing extended from at least February 28^th^, 2020, and in which the virus lineage was determined in all or a subset of the detected influenza B cases. We excluded articles in which: (a) the data collection period was entirely prior to 2020 or was not specified; (b) no respiratory specimens tested positive for influenza; (c) all influenza detections were due to type A viruses, or the virus type was not determined; (d) none of influenza B detections was tested to determine the virus lineage; or (e) there were no original data (e.g. commentaries, editorials, or letters without data). Articles that were entirely based on data from the WHO-FluNet, the Global Initiative on Sharing All Influenza Data (GISAID), or the Global Influenza Hospital Surveillance Network (GISHN) study were not considered, since an extensive search of these databases was also conducted as part of the present study (see below). We applied no restrictions in terms of study design: therefore, studies as diverse as surveillance reports, hospital-based clinical studies, vaccine-effectiveness studies, and phylogenetic analysis of influenza isolates (e.g. to determine their genetic and antigenic characteristics) were all considered as potentially eligible for inclusion as long as they matched the criteria listed above.

The data extracted from each eligible study included: study country, start and end of data collection, number of respiratory specimens that were collected and tested for influenza, number of those testing positive for influenza and specifically for type B influenza, and, among the type B influenza, the number of those that were characterized and the number that had a B/Yamagata lineage.

### Influenza B/Yamagata viruses reported in the WHO-FluNet, GIHSN, and GISAID

The FluNet (https://www.who.int/tools/flunet) is a web-based database that is coordinated by the WHO and is freely available to researchers. The FluNet database contains information on the weekly number of laboratory-confirmed influenza detections that are uploaded from WHO regional databases or directly entered by national influenza centres (NICs) and other influenza reference laboratories from 194 countries and territories (in August 2023) participating in the GISRS (Global Influenza Surveillance and Response System) initiative. Information on the influenza virus type is available for the vast majority of reported cases as well as the influenza A virus subtype, while the proportion of influenza B cases for which the lineage is reported varies considerably across countries and years [**Zanobini 2022**]. Influenza data from week 1 of 2020 onwards was downloaded from the FluNet on August 28^th^, 2023. The total number of influenza B/Yamagata detections and the number per 100,000 tested specimens, total influenza cases, and influenza B cases (overall and the subset of those that were characterized) were calculated for each of the following five periods: 2020 weeks 1-17 (January-April), 2020 weeks 18-53 (May-December), 2021, 2022, and 2023 weeks 1-33. Moreover, we collected the names of the countries in which influenza B/Yamagata detections occurred in each of these five periods.

The GIHSN (https://gihsn.org/) initiative was launched in 2012 through a partnership between industry, public health institutions, and sentinel hospitals in several countries around the world. The overarching objective of GIHSN is to collect data on the epidemiology of severe influenza, defined as a laboratory-confirmed influenza infection resulting in hospitalization, and provide annual estimates of the effectiveness of the influenza vaccine in preventing influenza-associated hospitalization. The GIHSN database was queried for severe influenza B/Yamagata cases reported from week 1 of 2020 to week 20 of 2023, with 21 countries contributing data on influenza in at least one season during this period (Brazil, Canada, China, France, India, Ivory Coast, Kenya, Lebanon, Mexico, Nepal, Pakistan, Peru, Romania, Russia, Senegal, Serbia, South Africa, Spain, Turkey, Ukraine, and the USA).

GISAID (Global Initiative on Sharing All Influenza Data; https://gisaid.org/) is a publicly accessible repository established in 2008 to share viral genomic data for influenza (later expanded to include SARS-CoV-2 and other pathogens). The GISAID database includes genetic sequences for all influenza viruses as well as related epidemiological data, and access is provided free-of-charge to users around the world in order to stimulate research and gain understanding on how influenza viruses evolve over time and spread globally. As GISAID includes influenza virus sequences from other sequence databases, such as GenBank, these other databases were not queried separately. The GISAID database was queried on 9 September 2023 for all B/Yamagata and B/unknown-lineage labelled sequences with a collection date from March 1^st^, 2020 onwards. Sequences were analysed using GISAID and GenBank blast and phylogenetic analysis using MEGA version 7.0.21 taking into account sequences of all available segments, and Nextclade version 2.14.1 (https://clades.nextstrain.org/) for hemagglutinin segment assessment.

### Review of national respiratory viruses surveillance systems’ webpages

During May and June 2023, we searched the web pages of influenza and respiratory viruses surveillance systems of the following countries: England, France, Germany, Italy, Netherlands, Portugal, Russia and Spain (Europe); Argentina, Brazil, Canada, Mexico and USA (Americas); China, Hong Kong, Israel, Japan and Turkey (Asia); Australia and New Zealand (Oceania); and South Africa (Africa). These countries were selected due to their geographical representativeness and availability of web-based surveillance reports. Extracted information was the number of respiratory specimens tested for influenza, as well as the number of influenza detections, overall and by virus type (A and B); if available for influenza B viruses, the virus lineage (Victoria, Yamagata, and not characterized) was also recorded. Moreover, we retrieved the available information on the main features of the surveillance system and the duration of the surveillance period (i.e. whether year-round or if limited to the usual epidemic period).

## Results

### Literature review

The literature search returned 2,147 entries in PubMed and 3,983 in EMBASE. The non-duplicate articles were 4,333, of which 3,746 were discarded based on the title and abstract (**Figure 1**). Of the remaining 587 articles, 547 were discarded for not matching the inclusion criteria (of these, 62 were not further considered since no reported influenza cases were type B), 7 for being based on the WHO FluNet and/or GISAID database, 3 for being previous reports from the GIHSN, and 4 because of overlap with more recent and/or larger articles based on the same data sources. A total of 26 articles were included in the systematic review: their main characteristics are reported in **Table 1** [**Soldevila 2022, Murillo-Zamora 2021, Korsun 2021, Panatto 2021, Pablo-Marcos 2020, Miron 2021, Hu 2021, Omer 2022, Olson 2022, Auvinen 2022, Kuzmanovska 2021, Wagatsuma 2022, Heinzinger 2021, Rios-Silva 2022, Suntronwong 2021, da Costa 2022, Melidou 2020, Huang 2022, O’Neill 2022, Kolosova 2022, Peck 2023, Chon 2023, Merced-Morales 2022, Melidou 2022, Sominina 2022, Song 2022**]. Europe was the most represented area (12 articles, of which 2 reported findings from multiple countries in the WHO European Region), followed by Asia (n=6), North America (n=5), and Oceania and South America (n=1 each). In addition, the paper by O’Neill et al. reported influenza positive samples coming from fourteen countries in Oceania, Asia, and Africa during 2020-2021, that were subtyped at the WHO CCRRI (Collaborating Centre for Reference and Research for Influenza) in Melbourne [**O’Neill 2022**].

**Figure 1.**
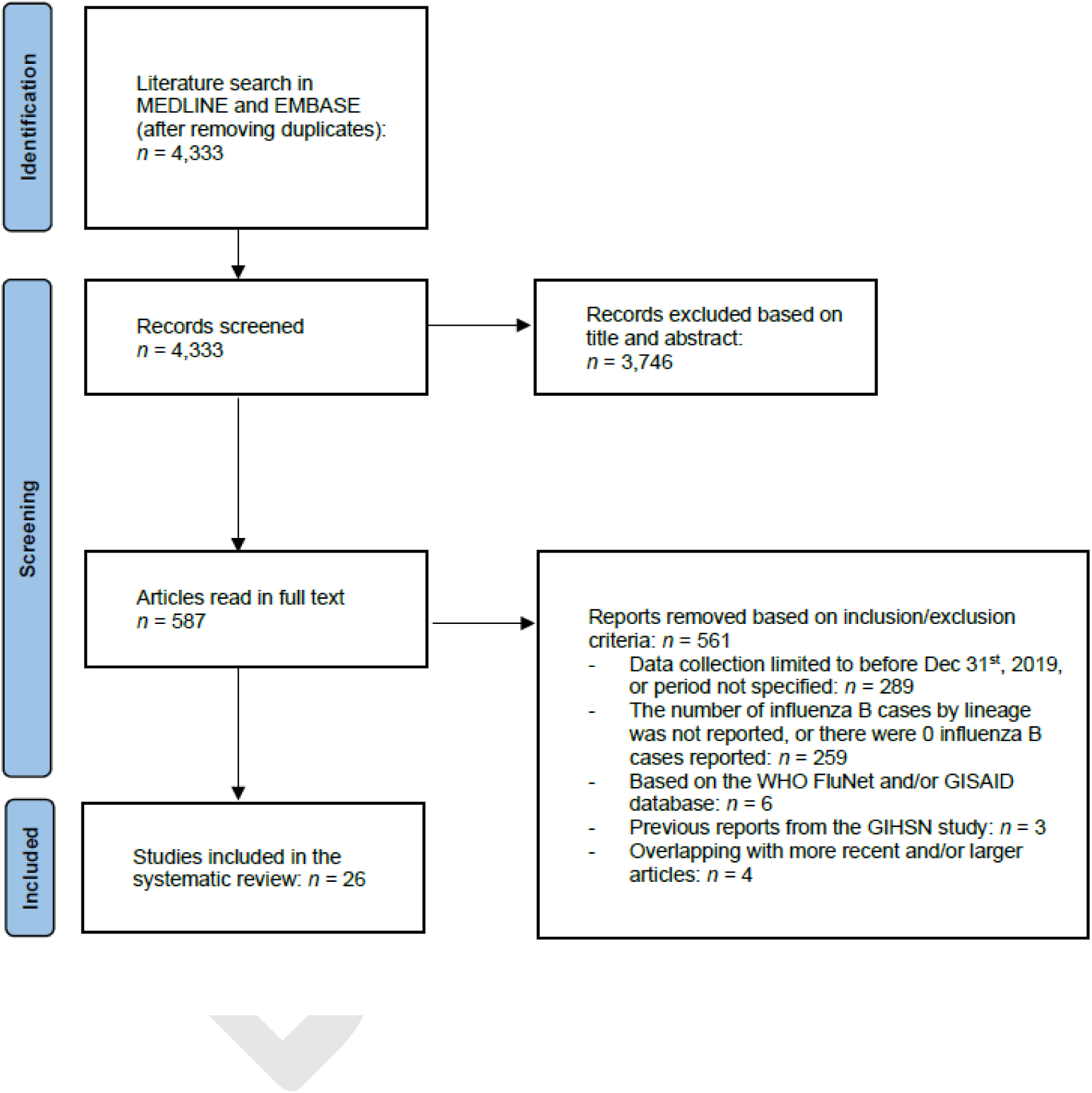
Flow-chart of the systematic literature review of articles reporting the number of influenza B/Yamagata virus detections from January 1^st^, 2020, until March 20^th^, 2023.

**Table 1.** Literature review of articles reporting on the number of influenza B/Yamagata virus detections from January 1^st^, 2020.

| Author, year | Country or world region | Start of observation period | End of observation period | N respiratory specimens | N influenza | N influenza B | N influenza B characterized | N influenza B Yamagata |
| --- | --- | --- | --- | --- | --- | --- | --- | --- |
| *Observation period from <u>before 2019</u> to <u>2020</u>* |  |  |  |  |  |  |  |  |
| Soldevila N et al, 2022 <sup>(a)</sup> | Spain | 2010, ns | 2020, ns | NA | NA | NA | 1,159 | 955 |
| Murillo-Zamora E et al, 2021 | Mexico | 2016, Oct | 2020, Apr | NA | 4,678 | 808 | 632 | 283 |
| *Observation period from <u>2019</u> to <u>2020</u>* |  |  |  |  |  |  |  |  |
| Korsun N et al, 2021 | Bulgaria | 2019, Oct | 2020, Mar | 1,387 | 610 | 138 | 138 | 0 |
| Panatto D et al, 2021 | Italy | 2019, Nov | 2020, Feb | 1,683 | 477 | 119 | 53 | 1 |
| Pablo-Marcos D et al, 2020 | Spain | 2019, Nov | 2020, Mar | NA | 652 | 180 | 167 | 0 |
| Miron VD et al, 2021 | Romania | 2019, Nov | 2020, Mar | 1,042 | 516 | 244 | 239 | 0 |
| Hu W et al, 2021 | USA | 2019, w45 | 2020, w13 | 28,176 | 5,529 | 2,336 | 862 | 6 |
| Omer I et al, 2022 | Israel | 2019, w46 | 2020, w11 | 1,769 | 973 | 354 | NA | 0 |
| Olson SM et al, 2022 | USA | 2019, Dec | 2020, Apr | 291 | 159 | 61 | 39 | 0 |
| Auvinen R et al, 2022 | Finland | 2019, Dec | 2020, Jun | 244 | 33 | 2 | 2 | 0 |
| Kuzmanovska M et al, 2021 <sup>(a)</sup> | Macedonia | 2019, ns | 2020, ns | NA | 251 | NA | NA | 0 |
| Wagatsuma K et al, 2022 <sup>(a)</sup> | Japan | 2019, ns | 2020, ns | NA | 159 | 53 | 53 | 0 |
| *Observation period from <u>2019</u> to <u>2021 or 2022</u>* |  |  |  |  |  |  |  |  |
| Heinzinger S et al, 2021 | Germany | 2019, w38 | 2020, w14 | 1,376 | 517 | 153 | 151 | 22 |
|  |  | 2020, w38 | 2021, w14 | 470 | 3 | 1 | 1 | 0 |
| Ríos-Silva M et al, 2022 | Mexico | 2019, Oct | 2020, Apr | NA | 4,836 | 1,910 | 1,843 | 92 |
|  |  | 2020, Oct | 2021, Apr | NA | 0 | - | - | - |
|  |  | 2021, Oct | 2022, Apr | NA | 816 | 7 | 4 | 0 |
| Observation period entirely in <u>2020</u> |  |  |  |  |  |  |  |  |
| Suntronwong N et al, 2021 | Thailand | 2020, Jan | 2020, Mar | NA | 349 | NA | NA | 0 |
| da Costa JC et al, 2022 | Brazil | 2020, Jan | 2020, Mar | NA | NA | 9 | 9 | 0 |
| Melidou A et al, 2020 | WHO Euro | 2020, w1 | 2020, w20 | NA | 1,691 | 397 | 397 | 1 |
| Observation period from <u>2020</u> to <u>2021 or 2022</u> |  |  |  |  |  |  |  |  |
| Huang WJ et al, 2022 | Northern China | 2020, Jan | 2020, Dec | 177,987 | 11,742 | 2,100 | 2,093 | 42 |
|  |  | 2021, Jan | 2021, Dec | 220,011 | 12,117 | 12,106 | 12,087 | 13 |
|  | Southern China | 2020, Jan | 2020, Dec | 294,656 | 14,698 | 6,817 | 6,719 | 15 |
|  |  | 2021, Jan | 2021, Dec | 336,851 | 25,227 | 25,163 | 24,966 | 13 |
| O'Neill G et al, 2022 | Melbourne WHO-CCRI | 2020, Jan | 2020, Dec | NA | 2,021 | NA | 65 | 2 |
|  |  | 2021, Jan | 2021, Dec | NA | 372 | NA | 13 | 0 |
| Kolosova NP et al, 2022 | Russia | 2020, Mar | 2021, May | NA | 6 | 6 | 6 | 0 |
|  |  | 2021, Jun | 2022, Feb | NA | 920 | 17 | 12 | 0 |

| *Observation period in <u>2021</u> or from <u>2021 to 2022</u>* |  |  |  |  |  |  |  |  |
| --- | --- | --- | --- | --- | --- | --- | --- | --- |
| Peck H et al, 2023 | Australia | 2021, Jan | 2021, Oct | 15,026 <sup>(b)</sup> | 42 | 10 | 10 | 0 |
| Chon I et al, 2023 | Myanmar | 2021, Oct | 2021, Dec | 414 | 16 | 3 | 3 | 0 |
| Merced-Morales A et al, 2022 | USA | 2021, Oct | 2022, Jun | 877,928 | 24,432 | 126 | 41 | 1 <sup>(c)</sup> |
| Melidou A et al, 2022 | WHO Euro | 2021, w40 | 2022, w10 | NA | 55,049 | 1,603 | 15 | 7 <sup>(d)</sup> |
| Sominina A et al, 2022 <sup>(a)</sup> | Russia | 2021, ns | 2022, ns | 137,161 | 8,637 | 275 | 275 | 0 |
| Song SX et al, 2022 <sup>(a)</sup> | China | 2021, ns | 2022, ns | 26,754 | 7,250 | 7,250 | 7,250 | 0 |

N: number of reported cases. WHO-CCRI: World Health Organization - Collaborating Centre for Reference and Research on Influenza
<sup>(a)</sup> The observation period was reported as the "season" (namely, the season 2019-2020 in Kuzmanovska et al. and Wagatsuma et al.; the season 2021-2022 in Sominina et al. and Song et al.; and the seasons 2010-2020 in Soldevila et al.).
<sup>(b)</sup> The specimens were collected from travellers arriving on repatriation flights..
<sup>(c)</sup> Other B/Yamagata detections were linked to LAIV, while only 1 was not tested.
<sup>(d)</sup> All reported 7 B/Yamagata were from Scotland and coincided with LAIV vaccination campaign, but further testing was not performed

In **Table 1**, studies were ordered according to the start and the end of the observation period. In two articles, data collection began several years before the start of the COVID-19 pandemic [**Soldevila 2022, Murillo-Zamora 2021**]: B/Yamagata viruses accounted for a large proportion of all characterized influenza B-positive specimens, but since data stratified by year were not available, it was not possible to assess how many of them were seen after March 1^st^, 2020. Ten studies reported data for the Northern hemisphere 2019-2020 influenza season: in only two of these there were B/Yamagata detections, which represented a very minor percentage of all influenza-positive specimens (1 out of 1,683 in Panatto et al, Italy [**Panatto 2021**], and 6 out of 28,176 in Hu et al., USA [**Hu 2021**]). In two more studies (Heinzinger et al., Germany, and Ríos-Silva et al., Mexico), data collection started in 2019 and ended in 2021 or, respectively, 2022 [**Heinzinger 2021, Rios-Silva 2021**]: in both studies, there were no B/Yamagata detections after the 2019-2020 season.

Three studies included data that were collected entirely in 2020: Melidou et al. analyzed European surveillance data from TESSy during the weeks 1-20 of 2020 and reported a single influenza B/Yamagata case (out of 1,691 influenza detections overall, of which 397 were influenza B) [**Melidou 2020**], while in the other two studies, conducted in Thailand and Brazil, the observation period was January-March 2020 and there were no reported B/Yamagata detections [**Suntronwong 2021, da Costa 2022**]. In nine more studies, the observation period started in 2020 or 2021, and ended in 2021 or 2022. In China, the proportion of influenza-positive specimens reported to the Chinese National Influenza Surveillance Systems that were caused by B/Yamagata lineage viruses was 0.22% in 2020 and 0.07% in 2021 [**Huang 2021**]. The remaining studies whose observation period started in 2020 or 2021 and ended in 2021 or 2022 reported no or a very limited number of influenza B/Yamagata detections, which after sequencing were found to be partly linked (in Melidou et al. 2022 and Merced-Morales et al. 2022 [**Melidou 2022, Merced-Morales 2022**]) to vaccination with LAIV.

In summary, we reviewed studies published up to March 2023 and whose observation period extended up to 2022, and found that B/Yamagata detections were reported only in a very few countries globally (and accounting for a very minor proportion of all influenza B cases) after the first few months of 2020.

### WHO-FluNet, GIHSN, and GISAID

On August 28^th^, 2023, we downloaded data from the *<u>WHO-FluNet</u>* for the period from week 1 of 2020 to week 33 of 2023. The database contained around 31.93 million respiratory specimens and 1,998,172 million influenza detections, of which 379,699 (19.0%) were influenza B viruses. Of these, 272,236 (71.7%) were not characterized, 107,048 (28.2%) were B/Victoria lineage, and 415 (0.1%) were B/Yamagata lineage. The average number of influenza B/Yamagata detections per 100,000 processed specimens dropped from 14.9 in the weeks 1-17 of 2020 to below 0.10 in 2022 and 2023 (**Table 2**). Influenza B/Yamagata accounted for a significantly greater proportion of detections during the weeks 18-52 of 2020 compared to the earlier weeks in the same year. This was followed by a major drop in the frequency of influenza B/Yamagata detections in 2021 and 2022 (**Table 2**): the number of influenza B/Yamagata detections dropped from 364 in 2020 to 45 in 2021 and three in 2022 (two from Germany and one from North Korea). In the first 33 weeks of 2023, there were 6 B/Yamagata detections globally, of which five were from Cameroon and one from Cuba: this represented a slight increase compared to 2022 but remained well below the numbers in 2020 and 2021 (**Table 2**). In summary, the rate of B/Yamagata detections entered in the WHO-FluNet database dropped significantly worldwide after 2020, both in absolute terms and as a percentage of the total number of influenza and influenza type B detections (**Table 2**).

**Table 2.** WHO-FluNet: number of influenza B Yamagata detections (overall and by country) in 2020 (weeks 1-17 or 18-53), 2021, 2022, and 2023 until week 19, and rate of detections per 100,000 processed specimens and influenza-, influenza B-, and characterized influenza B-positive detections.

| Year and period | Influenza B Yamagata detections | Influenza B Yamagata detections per 100,000: |  |  |  | Detections by country and week |
| --- | --- | --- | --- | --- | --- | --- |
|  |  | processed specimens <sup>(a)</sup> | influenza-positive detections | influenza B-positive detections | influenza B-positive detections, lineage characterized |  |
| 2020, weeks 1-17 | 351 | 14.88 | 75.27 | 211.54 | 1386.47 | **n = 117:** USA (weeks 1-13), **n = 67:** Mexico (weeks 1-14), **n = 54:** China (weeks 1-17), **n = 18:** Norway (weeks 1-8), **n = 14:** Dominican Republic (weeks 2-9), **n = 10:** France (weeks 2-12), **n < 10:** 24 countries |
| 2020, weeks 18-52 | 13 | 0.40 | 269.93 | 604.09 | 2245.25 | **n = 8:** USA (weeks 44-49), **n = 4:** China (weeks 19-51), **n = 1:** Afghanistan (week 45) |
| 2021 | 42 | 0.51 | 36.10 | 123.55 | 144.33 | **n = 32:** China (weeks 1-39), **n = 3:** India (week 41), **n = 1:** Afghanistan (week 1), Niger (week 1), Ghana (week 4), Bulgaria (week 8), Ivory Coast (week 8), Mexico (week 24), USA (week 39) |
| 2022 | 3 | 0.03 | 0.31 | 4.74 | 8.67 | **n = 2:** Germany (weeks 12-13), **n = 1:** North Korea (week 43) |
| 2023, weeks 1-33 | 6 | 0.09 | 1.40 | 5.25 | 33.61 | **n = 5:** Cameroon (weeks 23-31), **n = 1:** Cuba (week 11) |

<sup>(a)</sup> Calculated over the total of countries and weeks in which the number of processed specimens was reported.

The number of influenza detections within the *<u>GIHSN</u>* is reported in **Table 3**. Out of a total of 8,590 influenza detections reported between January 2020 and week 27 of 2023, 2,055 (23.9%) were influenza B. Of the 978 influenza B viruses that were characterized, seven (0.7%) were B/Yamagata, all detected in 2020 (two in France, two in Mexico, one in India and one in Russia in the first three months of the year, and again one in India in week 51). No B/Yamagata influenza detections were reported from 2021 onwards.

**Table 3.** Global Influenza Hospital Surveillance Network: number of influenza detections (overall, type B, characterized type B, and B/Yamagata) in 2020 (weeks 1-17 or 18-53), 2021, 2022, and 2023until week 29.

| Period | Influenza-positive detections | Influenza B-positive detections | Influenza B-positive detections, lineage characterized | Influenza B/Yamagata detections by country and week |
| --- | --- | --- | --- | --- |
| 2020, weeks 1-17 | 3,772 | 1,449 | 639 | 6 (France weeks 1 and 9, India week 5, Mexico weeks 6 and 9, Russia week 12) |
| 2020, weeks 18-53 | 15 | 5 | 1 | 1 (India week 51) |
| 2021 | 875 | 149 | 33 | 0 |
| 2022 | 2,961 | 241 | 173 | 0 |
| 2023, weeks 1-27 | 967 | 211 | 132 | 0 |

On September 9^th^, the *<u>GISAID</u>* database was queried for all influenza B/Yamagata and B/unknown-lineage sequences with collection date from March 1^st^, 2020, onwards. The set of B/Yamagata sequences included a total of 20 non-duplicate viruses. Collection dates were in March 2020 (n=14), November 2021 (n=4, all from Kenya), June 2022 (n=1, from Greece) and March 2023 (n=1, from the UK). Of these, two (from Japan, 2020, and Greece, 2022) had an HA genome segment identical to that of a B/Yamagata-like vaccine strain, suggesting both were vaccine-derived viruses. Of the five viruses from 2021 and 2023 only the PA (n=3) or MP (n=2) segment was available. For three of the four viruses from Kenya (2021), the limited available information made it difficult to determine whether they were from wildtype circulating B/Yamagata lineage viruses. For the remaining virus from Kenya and the virus from UK, the MP segment matched with highest rank with B/Victoria lineage viruses, suggesting that these detections were incorrectly labelled B/Yamagata. The B/unknown-lineage dataset included 128 unique viruses. The blast and phylogenetic analysis revealed that the majority (n=111) of these were wildtype B/Victoria and 7 were derived from the study published in 2023 by Aslam et al. on historic B viruses for influenza vaccine virus construction [**Aslam 2023**]. Of the remaining 10 viruses, two collected in the USA in 2021 and one from South Korea with data collection in 2020 were highly likely LAIV derived. Finally, seven waste water specimens collected in March 2020 in New York had partial PB1 sequences similar to 2007/2008 B/Yamagata lineage viruses, however, no sequences of other segments were available for definitive assessment. In conclusion, no sequences of wildtype viruses of the B/Yamagata lineage with a collection date after March 2020 have been uploaded to GISAID and GenBank, including the viruses reported as B/Yamagata lineage to FluNet and GIHSN after March 2020 (of note, therefore, for these later viruses there was no objective assessment possible whether these viruses were wildtype or vaccine-derived).

### Review of national respiratory viruses’ surveillance systems’ webpages

The number of influenza B/Yamagata detections reported by country and period is shown in **Table 4**, alongside with details of the surveillance system in which each case was observed and overall number of influenza cases reported. After week 35 of 2020, 11 B/Yamagata detections were reported: one in Australia (week 37-38 of 2020), nine in the USA (weeks 44-49 of 2021) and an additional case in the USA in week 46 of 2021. No B/Yamagata were reported during 2022 and 2023 until the time of our search in June 2023.

**Table 4.**
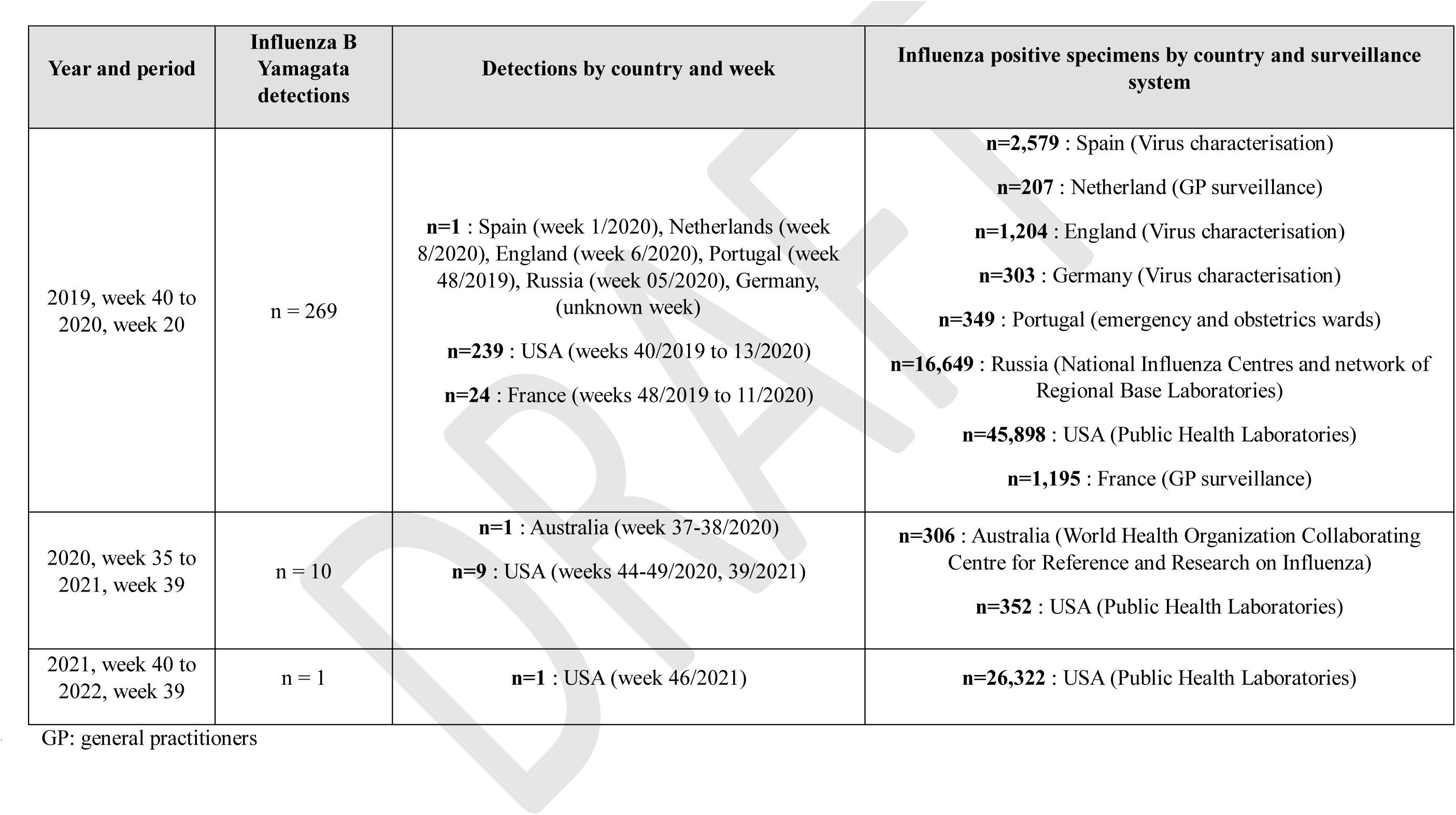
Number of respiratory specimens tested for influenza, and number of reported influenza B/Yamagata detections, from national influenza surveillance systems webpages worldwide (see text for a complete list of countries that were included).

To summarize the findings, we present the countries in which the last B/Yamagata detections (from any of the sources that we explored) was reported in 2023 (n = 2), 2022 (n = 3), or 2021 (n=9) in **Figure 2**. In all the remaining word countries, there we no B/Yamagata detections after 2020.

**Figure 2.**
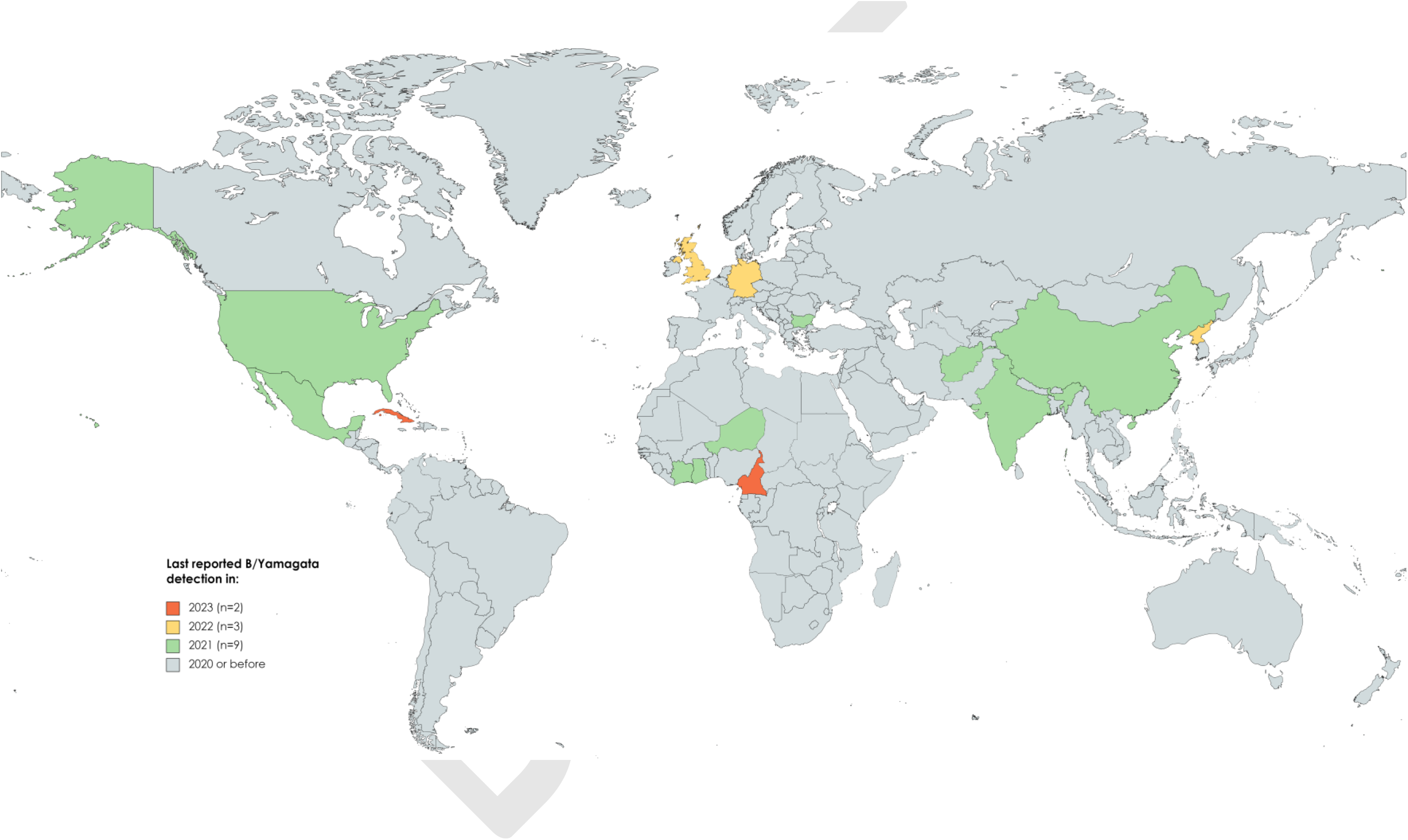
Map depicting the countries in which the last reported B/Yamagata detection occurred in 2023, 2022, 2021, or 2020 or before.

## Discussion

The early claims that the influenza B/Yamagata lineage may have gone extinct in the aftermath of the COVID-19 pandemic quickly raised the attention of policymakers (e.g. WHO) and vaccine manufacturers, as well as epidemiologists and researchers, due to the important implications this would have on the formulation and development of both inactivated and attenuated influenza vaccines. Given the enormous public health relevance and the need for solid and constantly updated epidemiological data, we undertook an extensive review of the available evidence on the topic, which covered multiple complementary sources of information and was therefore intended to be as comprehensive as possible.

Considering all of the collected information, the different sources investigated are concordant in showing that the number of influenza B/Yamagata viruses detections has declined considerably and progressively over the last 3.5 years (i.e. from the onset of the COVID-19 pandemic) compared with the pre-pandemic period, both in absolute terms (i.e. number of detections) and in terms of the positivity rate or as a fraction of the total number of influenza B detections. After the 2009 H1N1 influenza pandemic, influenza B viruses had resumed circulation causing about a quarter of all influenza cases globally on average [**Zanobini 2022**]. Following the major B/Yamagata outbreak that started in 2017 and involved large parts of the world, however, the circulation of B/Yamagata viruses had fallen considerably (they accounted for only one twentieth of all influenza B cases in 2018-2019 [**Zanobini 2022**]): this may help explain (together with the lack of known zoonotic or environmental reservoirs) why it is the only respiratory virus that has been pushed towards extinction by the emergence of SARS-CoV-2. Indeed, only a few detected B/Yamagata cases were reported in 2022-2023, but since the different world areas are unevenly covered by respiratory virus surveillance systems, some caution is warranted regarding whether the B/Yamagata lineage is actually extinct. However, the current epidemiological situation, with the rapid global disappearance (and potential extinction) of an influenza B virus lineage, is unprecedented since influenza B viruses split into two lineages in the early 1980s. In the 1990s, the B/Victoria lineage had practically disappeared from most world regions (including Europe and North America), but viral circulation endured in populous East Asia, so it was plausible to expect that it would re-emerge globally at some point (as it happened in 2001-2002) [**Xu 2015, Ikonen 2005**]. Today, the protracted and nearly undetectable viral circulation of B/Yamagata influenza viruses (including in East Asia) makes it much more likely that we are on the verge of B/Yamagata extinction, if it hasn’t already occurred. In this context, it is critical that B/Yamagata detections captured by surveillance systems or studies around the world within the context of research projects are properly investigated to determine whether B/Yamagata cases are wild-or vaccine-derived virus detections, and that a much larger proportion of influenza B detections are tested to determine their lineage. Vaccine-derived detections can mostly be linked to sampling LAIV vaccinated individuals and indeed, several B/Yamagata viruses whose sequences are uploaded in GISAID appeared to be vaccine-derived. It is therefore important sequencing of influenza B viruses is encouraged and supported and that procedures are put in place to assess the true circulation of the wild-type B/Yamagata virus, with detailed virological (sequence confirmation) and epidemiological (i.e. vaccination status, timing of vaccination and vaccine type) data being collected and shared (e.g. with the WHO and the WHO Collaborating Centres).

Aside from the question of whether B/Yamagata viruses have or have not already become extinct, the current epidemiological situation calls for a rapid response in terms of influenza prevention practices. In particular, a decision needs to be made regarding as to whether QIVs should be changed to TIVs and, if so, whether this change should be applied simultaneously to both egg-based vaccines and LAIVs. Any decision in this regard would have regulatory and manufacturing implications which would require different timescales to be implemented. Importantly from a public health perspective, it is critical that countries are not confronted with a situation where there are influenza vaccines shortages, thus these decisions would need to be taken with care and in a coordinated manner. Also, concerns exist about how the change from QIV to TIV should be communicated, as this could be confusing for the general public and have a negative impact on national influenza vaccination campaigns. Perhaps more theoretically (as implementation would be less immediate), the option of maintaining a quadrivalent vaccine by removing the B/Yamagata virus strain and replacing it with another strain (e.g. two H3N2 strains) would be worth discussing. Of note, there are other issues that are important beyond those concerning vaccine formulation and development. In particular, if B/Yamagata viruses are considered extinct, the question arises as to whether influenza laboratories around the world should change the security level with which they are allowed to handle B/Yamagata viruses, due to the potential risks associated with the escape of an extinct influenza virus.

A few aspects relating to the sources of information that we researched deserve comment. The WHO-FluNet database is dynamic in that the data it contains can change over time. For example, a NIC or reference laboratory in a given country may need to make corrections to data entered in previous weeks. As far as our current research question is concerned, it is worth noting that the WHO-FluNet database does not presently have an option to report vaccine-derived detections separately. We are aware that WHO proactively investigates all reports of B/Yamagata entered into their global surveillance databases and these are sometimes removed at a later stage when laboratory analyses reveal that they are vaccine-derived. In preparing this article, the 2020-2023 data from the WHO-FluNet database were downloaded twice, on May 19^th^ and then again on August 28^th^ 2023. In this short time period, the number of reported B/Yamagata influenza cases decreased from 45 to 42 for the year 2021, and from 8 to 3 for the year 2022. Hence, although the number of B/Yamagata detections in 2023, as shown in Table 2, slightly exceeded the number in 2022 (6 vs. 3), this might be an overestimate compared to reality (and the same considerations may apply to the GISAID database). A further limitation is the under-representation of tropical countries, particularly in populous South-East Asia and Africa, in data gathered from the review of scientific literature and webpages of respiratory viruses’ surveillance systems. This is important as we have previously shown that influenza B tended to circulate more intensely in the tropics than in the temperate countries before the COVID-19 pandemic [**Caini 2019**]. While a main strength of our data review is the systematic and comprehensive nature of the exercise, one should recognise the limitations and shortcomings of the information sources that we researched in order to be able to critically appraise the whole body of evidence currently available on this topic.

We have described the current epidemiological situation regarding B/Yamagata lineage viruses, and outlined its most important public health implications, particularly with regard to the composition of influenza vaccines. A further, and currently overlooked source of uncertainty with implications for immunization campaigns, relates to the timing of influenza epidemics in the coming years [**Caini 2018**]. Right now, there is in fact no guarantee that the pre-pandemic timing will be restored, or if instead, the co-circulation of SARS-CoV-2 will cause influenza epidemics to be shifted by a few weeks earlier or later due to viral interference [**Fage 2022, Piret 2022**], which might in turn entail the need to accommodate the timing of vaccine strains selection, manufacturing, and deployment. Furthermore, it is currently difficult to make predictions about the impact that the disappearance of B/Yamagata viruses will have on the circulation of other influenza viruses. B/Yamagata viruses tended to infect adults and the elderly more often than those of the B/Victoria lineage, which are to a much larger extent limited to children and adolescents [**Caini 2019**]. If B/Yamagata viruses actually became extinct, the age distribution of influenza B cases might shift towards younger ages, thus resulting in a lower disease burden than in the pre-COVID-19 era. However, this is currently nothing more than a plausible hypothesis that only time will allow one to confirm or refute. These many uncertainties imply that the role of epidemiological and virological monitoring of influenza viruses, SARS-CoV-2, and the other respiratory viruses in protecting the health of populations is presently more important than ever and will remain so for many years to come. In conclusion, international and national public health authorities (both public health institutes and medical regulatory authorities) are confronted with an unprecedented situation with the rapid disappearance and potential extinction of the influenza B/Yamagata virus (a development they have not experienced in the last 30 years) and need to take important decisions regarding influenza vaccines and the B/Yamagata virus, and we believe this review will contribute to this decision-making process.

## Data Availability

This is a review of data in the public domain.

## Acknowledgments

We would like to thank investigators at all sites participating to the Global Influenza Hospital Surveillance Network (GIHSN).

